# Preferences for Accessible Health Information Among Deaf, Blind, and DeafBlind Adults: A Descriptive Qualitative Study

**DOI:** 10.64898/2026.11.19.26368504

**Authors:** Logan S. Roberts, Christine Shu, Lisa Pizzo, PF Anderson, Dhruv Jain, Michael M. McKee, Tyler G. James

## Abstract

**Overview:** People with sensory disabilities (e.g., deaf or hard-of-hearing [DHH], blind/low vision [BLV], and DeafBlind) encounter difficulties when accessing health information. These barriers impede people with sensory disabilities in obtaining, understanding, and using health information. This study aimed to identify barriers, facilitators, and preferred formats for health information to inform inclusive health communication, while addressing the variety of accessibility needs across people who are DHH, BLV, and DeafBlind.

**Methods:** We conducted a qualitative study using eight focus groups and eleven individual interviews with adults with different types of sensory disabilities. Interviews were conducted directly in English or American Sign Language, and recordings were professionally transcribed. Discussions explored experiences accessing health information, information source preferences, accessibility barriers, and reactions to different formats. Transcripts were coded using a hybrid thematic analysis process. Themes were generated through team discussions, code synthesis, and community partner feedback.

**Results:** We developed three themes summarizing user-centered perspectives for designing consumer health information resources. Participants obtained health information from various sources including clinicians, family, friends, internet searches, and ChatGPT. Barriers included information overload, medical jargon, inaccessible website design, and communication breakdowns with clinicians. Preferences for health information formats differed across disability groups. Key recommendations included simplified language, structured layouts, and compatibility with assistive technologies (e.g., screen readers).

**Conclusion:** Despite varying accessibility needs across disabilities, participants agreed health information must be delivered through multiple accessible formats to meet diverse sensory needs. These findings are being used to develop health promotion checklists to enhance accessibility for people with sensory disabilities.

## INTRODUCTION

Health information plays a fundamental role in supporting informed decision-making, promoting healthy behaviors, and enabling individuals, including those with disabilities, to effectively manage their health and health conditions (Berkman et al., 2011; Rudwan et al., 2025; Robinson-Whelen et al., 2023). Effective, well-designed health information supports individuals in accessing, understanding, evaluating, and applying information to make informed decisions about their health (Berkman et al., 2011; Nam et al., 2023). However, these benefits depend on individuals being able to obtain, understand, and use health information that meets their communication and accessibility needs.

People with sensory disabilities – those who are deaf or hard-of-hearing (DHH), blind or low vision (BLV), and DeafBlind – frequently encounter barriers when accessing healthcare and health information. These barriers extend beyond the mere availability of or access to health education materials and include disability-related accessibility barriers (Agaronnik et al., 2019; Chandanabhumma et al., 2024; Chimienti et al., 2026; Cupples et al., 2012; James et al., 2022; Kim, 2019). For example, people with visual impairments may encounter difficulties obtaining health information in accessible formats (Cupples et al., 2012; Kim, 2019), while DHH individuals who use American Sign Language (ASL) may experience linguistic and communication barriers that affect access to health information (Chandanabhumma et al., 2024; McKee et al., 2015). Collectively, these barriers can limit an individual’s ability to obtain, understand, and use information needed to make decisions about their health and healthcare (Berkman et al., 2011; Nam et al., 2023; Robinson-Whelen et al., 2023).

Although increasing attention has been directed toward improving accessibility, meeting the diverse information and communication needs of people with sensory disabilities requires continued attention. Accessibility needs may differ across disability groups and among individuals within the same disability group, reflecting differences in communication preferences, functional needs, and approaches to accessing information (Agaronnik et al., 2019; Chandanabhumma et al., 2024; Choi & Chlebek, 2024; Cupples et al., 2012; Touchett et al., 2026). Consequently, a health information format that is accessible and usable for one individual or disability group may not adequately meet the needs of another.

Existing research has documented barriers experienced by people with disabilities when accessing healthcare and health information, including communication challenges during healthcare encounters, inaccessible information and materials, and inadequate accommodations (Agaronnik et al., 2019; Chandanabhumma et al., 2024; Chimienti et al., 2026; Cupples et al., 2012; Kim, 2019; McKee et al., 2015; Oswal & Oswal, 2021). Recent literature has also emphasized the importance of incorporating accessibility considerations and the perspectives of underrepresented users into the design of digital health resources (Smith et al., 2026). However, research examining user-centered preferences for accessible health information across multiple sensory disability communities remains limited, particularly as it pertains to digital information. Understanding both shared and unique preferences across DHH, BLV, and DeafBlind communities is critical for developing health communication strategies that are flexible, inclusive, and responsive to diverse accessibility needs.

The present study sought to identify barriers, facilitators, trusted information sources, and preferred formats for accessible health information among adults with sensory disabilities. Through focus groups and individual interviews with adults who are DHH, BLV, and DeafBlind, we explored how participants seek, evaluate, and use health information and identified recommendations for improving the accessibility of health communication. Findings from this study will inform the development of user-centered accessibility recommendations and health promotion resources designed to better meet the diverse needs of people with sensory disabilities.

## METHODS

### Study Design and Positionality

A qualitative design was selected to better understand participants’ lived experiences, perceived barriers, trusted health information sources, accessibility preferences, and recommendations for improving health communication. We conducted a descriptive qualitative study (Sandelowski, 2000; Sandelowski, 2010) using focus groups and individual semi-structured interviews to explore how adults with sensory disabilities access, evaluate, and use health information. Our research team brings extensive experience in disability and health research, accessible health communication, community-engaged research, public health, and qualitative research. Several team members have lived experience as family members of or people with sensory disabilities, bringing valuable perspectives that strengthen our work and help keep the needs and experiences of the disability community at the forefront. Team members vary in their personal and professional experiences with sensory disability and accessibility. which informed their perspectives throughout the research process. In addition to these experiences, the two principal investigators for this study are fluent in American Sign Language. We reflected on how these experiences along with our individual identities and disciplinary backgrounds influenced how we approach data collection, analysis, and interpretation throughout the research process. The study was approved by the University of Michigan Institutional Review Board (HUM00240346), and all participants provided informed consent prior to participation.

### Community Engagement

This study was guided by a user-centered, community-engaged approach supported by the project’s established Community Advisory Board (CAB) composed of individuals with lived experience of sensory disabilities, disability advocates, accessibility specialists, health literacy experts, librarians, researchers, and representatives from organizations serving DHH, BLV, and DeafBlind communities. Most CAB members represented multiple roles (e.g., a DHH librarian). Collectively, the CAB brought expertise in accessible health communication, patient education, assistive technology, disability advocacy, and information accessibility.

The CAB provided guidance throughout the research process. Prior to data collection, the CAB reviewed and refined the focus group and interview guides to ensure that discussion topics reflected community priorities, addressed issues most relevant to people with sensory disabilities, and were appropriate for participants with diverse communication needs. During data analysis, CAB members reviewed preliminary themes and provided feedback that informed the refinement of themes and subthemes, helping ensure that the findings accurately reflected participants’ lived experiences and perspectives. This collaborative approach strengthened the credibility, trustworthiness, and community relevance of the study (Lincoln & Guba, 1985) while informing the development of future accessibility recommendations.

### Participants and Recruitment

Adults 18 years of age or older who self-identified as DHH, BLV, or DeafBlind were eligible to participate. Participants were recruited across the United States through disability advocacy organizations, CAB partners, social media, and research networks affiliated with the University of Michigan. Recruitment aimed to include individuals across a range of demographic characteristics and lived experiences across the three sensory disability communities. Participants participated in the study through either a focus group or an individual interview and received $50 compensation for their time and participation.

### Data Collection

Data were collected between May and September 2025 through eight focus groups and eleven semi-structured individual interviews. The goal of the focus groups (60 to 90 minutes) were to facilitate discussion among participants with shared lived experiences, while interviews (30 to 60 minutes) were used to more deeply explore experiences and perspectives while increasing representation across sensory disability groups. Semi-structured interview and focus group guides were developed in collaboration with the research team and CAB partners to ensure that questions reflected the priorities and lived experiences of people with sensory disabilities. The guides included open-ended questions designed to explore participants’ experiences accessing health information, trusted health information sources, barriers to obtaining and understanding health information, preferences for accessible communication and information formats, and recommendations for improving the accessibility of health education materials. Participants also reviewed examples of health information materials and discussed design features that facilitated or hindered accessibility. Interviewers used follow-up questions and probes to encourage participants to elaborate on their experiences and provide examples.

Data collection sessions were facilitated by members of the research team with experience conducting qualitative and community-engaged research involving people with disabilities. Focus groups and interviews were conducted directly (i.e., without an interpreter) in either spoken English or American Sign Language (ASL), according to participant preference. Two focus groups and one individual interview were conducted in ASL, while the remaining sessions were conducted in spoken English. ASL sessions were video recorded, professionally translated and transcribed into English. English sessions were audio and video recorded and professionally transcribed. Members of the research team reviewed all transcripts for accuracy prior to analysis.

### Data Analysis

Data were analyzed using a hybrid thematic analysis approach that incorporated both deductive and inductive coding (DeJonckheere et al., 2024; Swain, 2018). An initial coding framework was developed based on the study objectives and the focus group and interview guides. Additional codes were developed throughout the analysis to capture concepts that emerged directly from participants’ experiences. Transcripts were managed and coded using MAXQDA software (Version 26.1.0). Members of the research team independently coded transcripts and met to compare coding, refine code definitions, and resolve discrepancies through discussion and consensus. Codes were iteratively grouped into categories and synthesized into broader themes and subthemes that reflected recurring patterns across participant experiences. Throughout the analysis, the research team compared findings across DHH, BLV, and DeafBlind participants to identify both shared experiences and differences in accessibility preferences. Themes were refined through iterative discussions among the research team and feedback from CAB partners to ensure that the findings accurately represented participants’ perspectives.

### Rigor and Trustworthiness

Several strategies were used to enhance the rigor and trustworthiness of the findings (Fereday & Muir-Cochrane, 2006). First, the study included participants from three sensory disability communities and incorporated both focus groups and individual interviews, allowing findings to be compared across participant groups and triangulated across data collection methods. Second, multiple members of the research team participated in coding and theme development using an iterative consensus process to strengthen analytic credibility. Third, peer debriefing was conducted with colleagues in the University of Michigan Mixed Methods Program (University of Michigan Mixed Methods Program Core Facility, n.d.) who were not directly involved in the study, providing an impartial review of the study methods, data, and interpretations. Fourth, CAB partners reviewed preliminary findings and provided feedback during theme refinement, helping ensure that the findings reflected participants’ lived experiences and priorities. Finally, the use of accessible data collection modalities (including data collected in the preferred language of participants) help increase representation of groups frequently ignored in mainstream public health research.

## RESULTS

### Participant Characteristics

A total of 30 adults with sensory disabilities participated in this study; all participants self-identified as deaf or hard of hearing (DHH), blind or low vision (BLV), and DeafBlind. Participants were in either one of eight focus groups, or one of eleven individual semi-structured interviews. Demographic characteristics of the study sample, including age, race and ethnicity, level of severity, and frequency of internet use for health information, are presented in **Table 1**. Participant characteristics and data collection methods are summarized in **Table 2**.

**Table 1.** Participant Demographics and Health Information Characteristics.

| **Characteristic** | **Overall Sample (N = 30)** |
|---|---|
| Age (years) |  |
| 18-24 | 3 |
| 25-34 | 3 |
| 35-44 | 5 |
| 45-54 | 6 |
| 55-64 | 4 |
| 65+ | 9 |
| Race/Ethnicity |  |
| White | 18 |
| Black or African American | 5 |
| Multiracial | 5 |
| Asian | 1 |
| Other | 1 |
| Sensory Disability and Self-Reported Severity |  |
| Deaf or hard-of-hearing |  |
| Mild/Moderate | 6 |
| Severe | 5 |
| Blind or low vision |  |
| Mild/Moderate | 6 |
| Severe | 3 |
| Not Reported | 1 |
| DeafBlind |  |
| Mild/Moderate | 6 |
| Severe | 3 |
| Frequency of Using the Internet for Health Information |  |
| Rarely | 0 |
| Sometimes | 9 |
| Often | 10 |
| Every day or frequently | 10 |
| Not Reported | 1 |

*Note. “Multiracial ” includes participants who selected more than one race and/or ethnicity, including participants who identified as white and Hispanic. Counts for individual categories reflect participants who selected that category alone.*

**Table 2.** Participant characteristics by sensory disability group and data collection method.

| **Disability Group** | **Focus Groups Participants** | **Interviews Participants** | **Total Participants** |
|---|---|---|---|
| **Deaf or hard-of-hearing** | 6 | 5 | 11 |
| **Blind or low vision** | 8 | 3 | 11 |
| **DeafBlind** | 5 | 3 | 8 |
| **Total** | **19** | **11** | **30** |

### Key Thematic Findings

Three overarching themes were identified across focus groups and individual interviews: (1) trust and credibility shape how individuals with sensory disabilities seek and evaluate health information; (2) accessible design and presentation of health information are essential for people with sensory disabilities to access and understand information; and (3) health information must be delivered in multiple accessible formats to meet diverse sensory needs. An overview of the three themes is presented in **Figure 1**.

**Figure 1.**
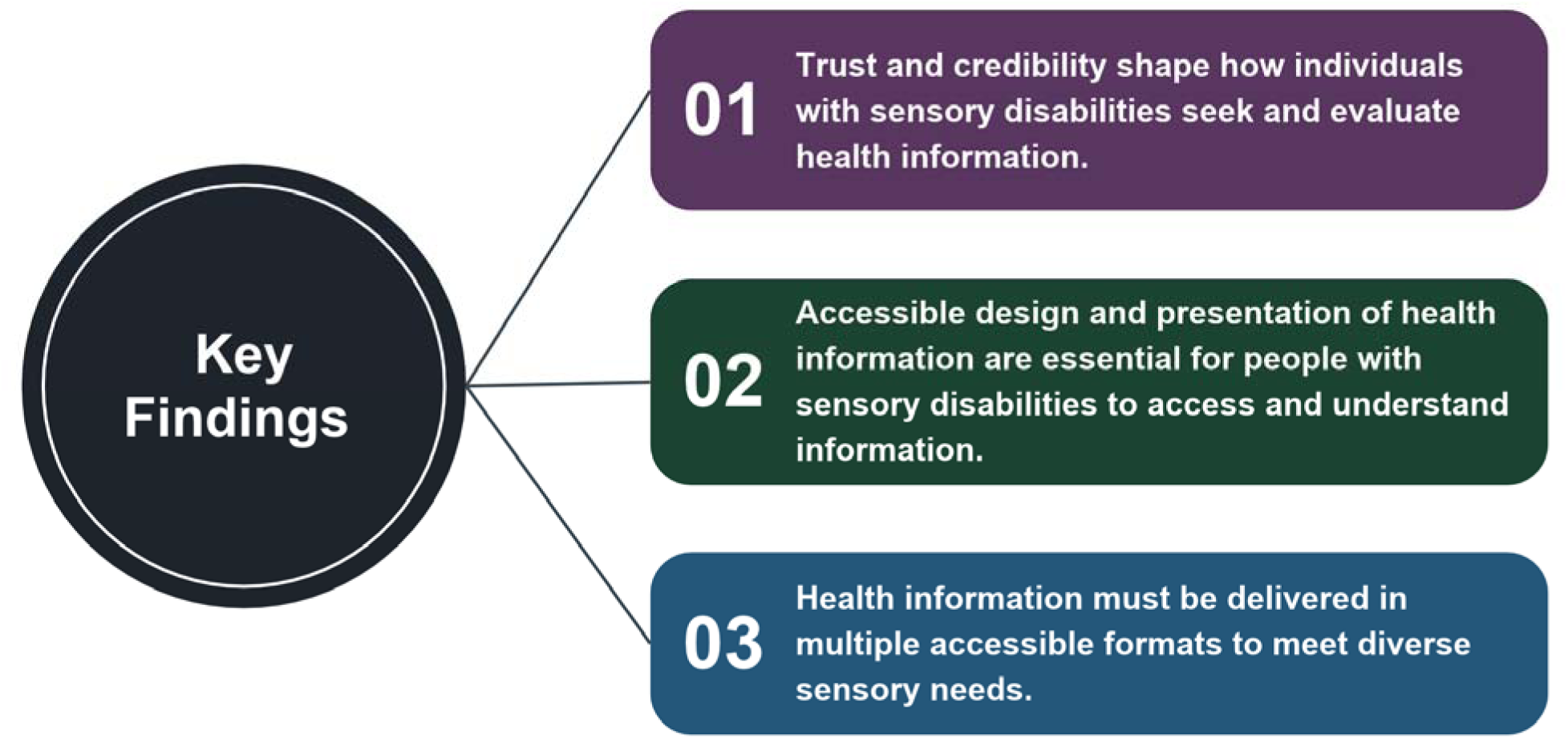
Overview of Key Thematic Findings

## Theme 1. Trust and credibility shape how individuals with sensory disabilities seek and evaluate health information

Participants described trust as the foundation of appraising health information resources. Regardless of disability group, participants emphasized the importance of obtaining information from sources they perceived as credible before making health decisions. Although many participants reported beginning their search online, few relied on single sources of information. Instead, participants described evaluating credibility of information by consulting healthcare providers, established medical organizations, and trusted family members or peers. Participants also acknowledged using emerging technologies, such as ChatGPT, and social media to obtain health information, although these sources were generally viewed as supplemental rather than definitive. Preferred health information sources reported by participants across sensory disability groups is summarized in **Table 3**.

**Table 3.** Preferred health information sources reported by participants with sensory disabilities.

| Health Information Source | Number of Mentions (n) |
|---|---|
| Healthcare providers (e.g., doctors, pharmacists) | 23 |
| Internet/Google search | 20 |
| Mayo Clinic | 10 |
| Cleveland Clinic | 9 |
| Friends and family | 8 |
| Personal testimonies (e.g., peer experiences and personal health stories shared online or in person) | 7 |
| Patient portals | 6 |
| WebMD | 6 |
| Social media (e.g., social media platforms used for medical or health information) | 6 |
| ChatGPT/AI | 5 |
| Other (e.g., NIH, medical journals) | 5 |

*Note. Frequencies represent the number of times participants mentioned each health information source across all focus groups and interviews. Each source was counted only once per participant, regardless of how many times the participant mentioned that source. Participants often reported using multiple sources when seeking health and therefore could be counted in more than one category.*

Trust was shaped by several factors, including the perceived expertise and credibility of the source, prior positive experiences using health information from the source, recommendations from healthcare professionals, and opportunities to verify information across multiple sources. Participants frequently described cross-checking information before acting on it, reflecting an intentional process of evaluating credibility rather than passively accepting health information.

### 1.1. Trusted health information sources

Healthcare providers were consistently described as the most trusted source of health information. Participants valued physicians, pharmacists, and other healthcare professionals because they were perceived as knowledgeable, familiar with participants’ medical histories, and able to provide individualized recommendations.

> “I typically go to my doctor because they’re the ones that diagnosed me.” (BLV participant)
>
> “…there’s an abundance of stuff out there and it’s like I don’t know the credibility of whoever is putting out there. So I just go to the source. Go right to the physician or the pharmacist.” (DHH participant)

Several participants explained that although they searched for information independently, they ultimately relied on conversations with their healthcare providers to confirm or clarify what they had learned.

> “I’m a Google person. I look for all my information online and then I ask doctors. I find out everything I can…I come informed and then I, because I don’t trust everything online, I reevaluate it, talk with my doctor about it and see what they have to say.” (DHH participant)

Participants also expressed strong confidence in established medical organizations, including the Mayo Clinic, Cleveland Clinic, National Institutes of Health (NIH), Centers for Disease Control and Prevention (CDC), and disease-specific organizations. These sources were viewed as evidence-based and more reliable than general internet searches. Rather than relying on the first result returned by a search engine, participants often described comparing information across multiple reputable organizations before drawing conclusions.

> “I’ve read up usually using the internet sites, sticking to sites that are trustworthy: the major clinics, Cleveland, Mayo…” (DeafBlind participant)
>
> “…Once I get the sources, I look through National Library of Medicine, and pubmed.gov…” (DHH participant)

For many participants, trusted information sources extended beyond healthcare professionals and medical institutions. Family members, friends, and peers with healthcare expertise or lived experience also played an important role in supporting health information seeking. Participants frequently described consulting relatives who worked in healthcare or relying on trusted members of their community to help interpret or explain complex medical information. For some BLV participants, family members also served as an important accessibility resource by reading or relaying health information that was difficult to access independently.

> “…having low vision, and the struggle of trying to get information, most times I rely on my nephew and nieces…They relay it back to me” (BLV participant)
>
> “I tend to ask my sister who is a doctor. She can sign [ASL]. I will ask what this means and she will explain to me…” (DeafBlind participant)

### 1.2. Supplementary and experiential information sources

Although healthcare professionals and established medical organizations were considered the most trustworthy sources of health information, participants also described using a variety of supplementary information sources to answer questions, gather personal experiences, or identify topics for further discussion with their healthcare providers. These sources included Google searches, ChatGPT, patient portals, social media platforms, online discussion forums, and personal testimonies shared by individuals with similar health conditions.

Participants acknowledged that social media and personal narratives from individuals with similar experiences or medical conditions could provide valuable insights into lived experiences, particularly when navigating chronic conditions, considering treatment options, or learning from others’ experiences with care.

> “Sometimes you learn from other people sharing their experiences, sharing stories.” (DHH participant)
>
> “I also found it really helpful to join some Facebook groups on sciatica or vision problems or medication issues or something like that. Then you get the experience from people who have tried and true solutions or sometimes are just as frustrated as you.” (BLV participant)

However, participants also recognized limitations of these sources and emphasized the importance of confirming information through trusted healthcare professionals or reputable medical organizations before making decisions.

> “I think Facebook is absolutely great… it’s a wealth of information… But… people also have to take it as a grain of salt, that it might work for me but not you.” (BLV participant)

Similarly, participants viewed artificial intelligence tools such as ChatGPT as useful for obtaining quick summaries or explanations of health topics but generally expressed caution about relying on AI as a sole source of medical information. Instead, participants described using AI as a starting point that prompted additional verification through trusted sources. Participants consistently distinguished between identifying information and trusting information.

> “I clicked page after page of the links, and after a while, I wasn’t satisfied. So, I went to ChatGPT and it gave me specific information on what I was looking for…I wasn’t satisfied, so I did ask for source links, and clicked on them. It helped.” (DHH participant)

Overall, participants demonstrated a thoughtful and deliberate approach to health information seeking. Rather than relying exclusively on any single source, they combined information from healthcare providers, reputable medical organizations, family members, peers, internet searches, and emerging technologies to develop a more complete understanding of their health concerns.

## Theme 2. Accessible design and presentation of health information are essential for people with sensory disabilities to access and understand information

Although participants emphasized the importance of obtaining health information from trusted sources (see Theme 1), they consistently described that credibility alone was insufficient if information was not accessible or easy to understand. Across all three sensory disability groups, participants identified design features that either facilitated or hindered their ability to engage with health information. Participants described accessibility as multidimensional, reflecting the interaction of visual, structural, and cognitive design features rather than any single accommodation. Even highly credible health information became less useful when it was difficult to read, poorly organized, or written using overly technical language. Participants emphasized that accessible design should minimize unnecessary barriers, support comprehension, and facilitate independent access to health information.

Participants further emphasized that accessibility was highly individualized. Features that improved accessibility for one person did not necessarily meet the needs of another, and no single format was viewed as sufficient for every individual. These perspectives reinforced the importance of considering diverse sensory abilities, communication preferences, and assistive technology use when designing health information and providing multiple accessible options to meet varied needs.

### 2.1. Visual accessibility

Participants consistently emphasized that visual presentation played a critical role in determining whether health information was usable. Font size, font style, color contrast, spacing, captions, and other visual communication supports were frequently discussed as factors that either supported or limited access to health information. Rather than viewing these features as aesthetic preference, participants described them as essential components of accessibility that directly influenced their ability to independently obtain health information.

BLV participants frequently described difficulties reading websites because of small fonts, poor color contrast, cluttered backgrounds, and limited magnification options.

> “Basically, information that’s health related and that usually I’m having difficulties in is font type. Different font type[s], magnification issues. The background color of the display pages. Most times the gray, the blue gray background color of most [landing] pages actually I find it difficult to read text from it… regular popups that comes along with it could be very, very disturbing too.” (BLV participant)

In contrast, participants responded positively to examples of health materials that incorporated larger fonts, adequate spacing, and readable color contrast.

> “The bold fonts, the font text size are bold… it’s just so clear. And the words aren’t jam-packed together, just properly spaced.” (BLV participant)
>
> “Blue or black is usually the easiest for me to see and having the white on the blue background…” (DeafBlind participant)

Participants also responded positively to the use of visual elements, such as infographics, that presented health information in an engaging and accessible way.

> “[Infographics] It draws my eye… somebody has a language barrier… they can look at this and see the little tidbits with some color… without having to read the boring text.” (Deafblind participant)

Participants who were DHH also emphasized the importance of visually accessible alternatives to spoken information in video-based health resources, including captions and sign language. These features provided access to information that might otherwise be communicated primarily through audio. Captioning was viewed not simply as an accommodation but as an important component of access to health information.

> “If it goes into detail, I prefer a spoken English video with captions.” (DHH participant)
>
> “It was very clear to understand what was going on… Having the captions certainly helped clarify what the couple were saying. I would say that was a pretty good format, having the captions, having the audio, for me was helpful.” (DeafBlind participant)
>
> “I wish there was a signer with subtitles in every video. I think it would be very good for the people who are hard of hearing like me that use sign language.” (DHH participant)

Collectively, these findings demonstrate that visual accessibility involves much more than increasing font size. Participants described visual accessibility as the combination of readable text, thoughtful use of color and spacing, captioning, magnification options, and visual elements that support independent engagement with health information.

### 2.2. Structural accessibility

Beyond visual features, participants emphasized the importance of how health information was organized, presented, and navigated. Clear navigation, logical organization, and well-structured pages were described as making it easier to locate relevant health information and navigate resources independently.

Several participants explained that difficult-to-navigate webpage layouts made it difficult to locate relevant information. They described abandoning websites that were poorly organized or contained excessive advertisements, pop-ups, or other distracting elements because finding relevant information required considerable effort.

> “The immense overload of distractions on all sorts of web pages: advertisements, popups… I like clean, simple graphics… I’m annoyed by things that distract me.” (DeafBlind participant)

Participants appreciated health resources that clearly separated information into manageable sections using headings, spacing, and other organizational features. These features made it easier to locate important information and navigate health resources independently.

> “…a medical website like that, that is just so straightforward, able to access the text; able to navigate… having a clean website like that, it’s going to really be good.” (BLV participant)
>
> “The headers are really big and bold, and I can tell from the header if the paragraph’s worth reading, which is nice cause then I can just get the information I really need and skip to the next thing much faster.” (DeafBlind participant)

Participants also described how compatibility with assistive technologies, including screen readers, text-to-speech software, and screen magnification, determined whether digital health information could be accessed independently.

> “If you try and enable the built-in web-based screen reader, and leave your existing screen reader enabled, it totally screws everything up, because they’re basically conflicting with each other…If it’s not functional, it really doesn’t do us any good.” (BLV participant)
>
> “If it’s a website… that’s not accessible to a screen reader user at all unless they use some kind of an alt text or something.” (BLV participant)

One participant described text-to-speech as particularly helpful, explaining, “I can just highlight it, and it will read it. It will read whatever it is for me.” (BLV participant). When websites were incompatible with these technologies, participants often relied on family members or others to assist with obtaining health information.

Participants consistently emphasized that the organization of information was important regardless of format. Whether information was presented through text, graphics, or multimedia, participants valued resources with clear headings, logically arranged sections, adequate spacing, and uncluttered layouts that made information easier to locate and navigate.

Overall, participants described structural accessibility as supporting their ability to independently locate, navigate, and access health information through clear organization, intuitive navigation, and compatibility with assistive technologies.

### 2.3. Cognitive accessibility

Participants also emphasized that accessible health information must be understandable. Regardless of disability group, participants frequently described challenges associated with medical jargon, lengthy explanations, dense text, and information overload. Accessible information was consistently described as information that communicated complex health concepts using clear, concise, and easy-to-understand language without sacrificing accuracy.

Participants noted that medical terminology often created unnecessary barriers, particularly when unfamiliar terms required additional searching before the original health information could be understood. Rather than facilitating learning, excessive jargon increased frustration and interrupted participants’ ability to engage with health information.

> “It’s hard to read. Also because of the technical terms, a lot of technical terms, there’s medical terms. Those are really stressful. Going to have to always go back to… Google… maybe it’s a medical word, and I have to also do my research about the word. I think that’s where the frustration comes in.” (BLV participant)
>
> “If I feel the information is clear with minimal jargon and is easy to understand, with images. If it’s too wordy or has a lot of terminology, then no. I weed through the contents to find simpler, explanatory sites…” (DHH participant)

Participants also described becoming overwhelmed when presented with large amounts of information at once. Long webpages, densely written educational materials, and excessive detail often discouraged continued reading.

> “I would much rather have short, concise stuff versus… lengthy explanations on things… Because then it just gets overwhelming.” (DHH participant)
>
> “The most important things go first. Because people get overloaded, and they set stuff down. Most of the things that people get sent, they read the first part cause they’re told they need to. And then they get overwhelmed, and they set it down.” (DHH participant)

To reduce cognitive burden, participants agreed on using plain language, concise explanations, and prioritizing key information. Many participants preferred resources that communicated key messages first, while providing opportunities to access more detailed information if desired.

Collectively, these findings demonstrate that cognitive accessibility extends beyond reading level alone. Participants emphasized that information should be organized and written in ways that reduce cognitive burden, allowing individuals to efficiently locate, understand, and apply health information. Together, the visual, structural, and cognitive accessibility needs described in Theme 2 highlight the importance of designing health information that is responsive to diverse sensory needs and preferences. Theme 3 further explores how participants described the importance of delivering health information through multiple accessible formats.

## Theme 3. Health information must be delivered in multiple accessible formats to meet diverse sensory needs

Although participants identified design features that improved accessibility, they also emphasized that no single communication format could meet the needs of every person with a sensory disability. Rather than advocating for one universally accessible resource, participants described accessibility as the availability of multiple formats that allow individuals to select the option that best aligns with their sensory abilities, communication preferences, health literacy, and use of assistive technologies. Importantly, participants explained that accessibility preferences varied not only across DHH, BLV, and DeafBlind communities, but also among individuals within the same disability group. Even within disability-specific focus groups, participants considered the accessibility needs of people with sensory disabilities other than their own when discussing how health information should be delivered. These findings suggest that accessible health information should prioritize flexibility and user choice rather than relying on a one-size-fits-all approach.

### 3.1. Accessibility needs vary across and within sensory disability groups

Participants recognized that accessibility needs differed across and within sensory disability communities. Although many recommendations were shared across participant groups, preferences for communication methods, educational materials, and information delivery varied depending on participants’ vision and hearing, communication preferences, previous experiences, and familiarity with assistive technologies. Participants frequently cautioned against assuming a single accessibility feature would adequately address the needs of all individuals. Participants also highlighted the importance of involving people with sensory disabilities in the development of accessible health information resources to ensure that their individual needs and preferences are considered throughout the development process.

> “It’s important to accommodate [a person’s] preferences, because not everyone is the same…disabled people have different needs and accommodations. Ask them and be willing to listen, and give them different options, whether it’s VRI, reading English, or having an onsite interpreter…The important thing is being open-minded and being willing to fit their needs.” (DHH participant)
>
> “I think that it’s important to acknowledge and provide resources for people with different abilities. I think it’s taking into consideration when commercials or advertisements… there are some people that are hearing impaired, some that are visually impaired, and tailor the products for those individuals.” (BLV participant)

Several DHH participants explained that written English may not be the preferred method of receiving health information. ASL is a distinct language with its own grammar and structure; written English is not equally accessible for all Deaf individuals.

> “…not all Deaf people understand English…ASL is a visual language, not written.” (DHH participant)
>
> “Some might find signing easier to understand, while others might struggle with English. So provide a signed version for all people.” (DeafBlind participant)

Similarly, participants who were BLV or DeafBlind emphasized the importance of Braille, screen readers, audio descriptions, and other features that support independent access to written information.

> “There should be more of Braille information, health information… I’m actually very good with reading Braille… There should be more information released in Braille.” (BLV participant)
>
> “Some can use screen readers or audio descriptions. For fully deaf and fully blind people, are all medical information accessible in Braille or transcript format? No. They don’t have full access.” (DeafBlind participant)

Participants also highlighted that preferences could vary even within the same individual depending on the situation. Some participants preferred reading text-based resources when seeking information quickly, but preferred videos when learning more complex health topics. Others described selecting different formats depending on their purpose for seeking information and the accessibility features available. For example, participants frequently used text-based resources to quickly locate or review information but preferred videos when they wanted more detailed explanations or step-by-step demonstrations.

> “When I finish Google, I will go over to YouTube and watch videos, which have everything: text, images, and action.” (DHH participant)

These differences in format preferences reinforce the importance of offering multiple accessible options rather than relying on a single communication format. Providing multiple options allows individuals to select the format that best meets their needs and preferences at a particular time.

### 3.2. Multimodal information formats

Participants recommended that health information be delivered through multiple complementary formats rather than relying on a single educational resource. Across disability groups, participants described value in providing options such as written materials, videos, graphics, captions, audio descriptions, Braille, and plain-language text, along with compatibility with assistive technologies to increase the likelihood that health information could be accessed independently.

> “Now, I’ve never seen anywhere where I have been presented with what I call ‘multiple modalities of communication,’ so a blind person, a hard of hearing-deaf person…a low-vision person. So, I think in theory it sounds like a great idea to have that same information communicated in multiple, different modalities.” (BLV participant)
>
> “Some people are more audio, some people are more visual learning. I think the more people that you can reach with whatever you’re using, the better.” (DeafBlind participant)

Participants also provided examples of how different communication formats could be made available alongside one another, allowing individuals to access the same information in their preferred format.

> “…you have someone sign in ASL what that form means, what it includes, and what is required. All of that while also providing a transcript for those who prefer to read, or an infographic. Provide this information both in English and ASL. I think that’d be best.” (DeafBlind participant)

Participants also described multimodal communication as promoting independence. When information was available in several accessible formats, individuals could choose the option that best supported their communication preferences without relying on family members, friends, or healthcare providers to interpret or explain information. Participants noted that this flexibility was particularly important because accessibility needs often changed depending on the health topic, setting, or technology available.

> “It’s health information needed for the general public. So, I feel there should be an option to toggle between deaf…blind…you can toggle between the information that you want, based on your sensory…sensory disability.” (BLV participant)

Importantly, participants clarified that multimodal communication did not require every resource to include every accessibility feature. Rather, they emphasized that health information should exist within an ecosystem of complementary resources that collectively meet the diverse needs of people with sensory disabilities. Participants preferred having meaningful choices among accessible formats over expecting a single resource to serve every possible communication need.

These findings highlight that accessibility is not achieved through standardization alone. Instead, participants envisioned a flexible system in which health information is intentionally developed in multiple accessible formats, allowing individuals to select the communication method that best aligns with their needs, preferences, and circumstances.

## DISCUSSION

This study explored how adults with sensory disabilities seek, evaluate, and use health information and identified user-centered recommendations for improving the accessibility of health communication. Across DHH, BLV, and DeafBlind communities, participants described health information seeking as a deliberate process shaped by trust, accessibility, communication, and individual preferences. Although participants identified many shared recommendations for improving accessibility, they consistently emphasized that no single communication strategy or format could meet the diverse needs of all individuals with sensory disabilities. Instead, participants described accessible health information as information that is trustworthy, intentionally designed, communicated through multiple accessible formats, and responsive to individual communication preferences.

A primary finding from this study is that participants viewed accessibility as extending far beyond individual accommodations or compliance with accessibility standards. Discussions of accessibility frequently included visual design, page organization, plain language, compatibility with assistive technologies, communication supports, and flexibility in information delivery. These findings are consistent with previous research documenting barriers related to inaccessible or unavailable alternative formats, difficulty reading and understanding health information, and limitations in the usability of assistive technologies among people with visual disabilities (Kim, 2019). Recent research with blind and visually impaired adults has similarly emphasized the importance of simplicity and customizable design features, including adaptable visual and auditory presentation of information (Choi & Chlebek, 2024). Similar barriers to independent digital health information access have been documented among blind and low-vision users, including inaccessible websites, data visualizations, and digital platforms and incompatibility with screen readers (Oswal & Oswal, 2021).

Rather than focusing on isolated accessibility features, participants described accessibility as an integrated process that supports independent access to health information. In particular, structural accessibility, including clear organization, intuitive navigation, and compatibility with assistive technologies, may support greater independence by reducing barriers to locating and accessing health information. These findings suggest that improving health communication requires consideration of how individuals locate, navigate, understand, and apply health information throughout their healthcare experiences.

A second finding was participants’ emphasis on flexibility rather than standardization. Although many accessibility guidelines focus on implementing specific accommodations, participants consistently described that accessibility preferences differed across and within sensory disability groups. This is consistent with previous research among DHH adults demonstrating differences in health information support needs and approaches to accessing and using information, including differences associated with health literacy (Chandanabhumma et al., 2024). Chandanabhumma et al. (2024) also found that DHH participants valued tailored information dissemination approaches, including textual accommodations and ASL-based information.

For example, some participants preferred written materials, while others favored videos, Braille resources, captions, audio descriptions, or American Sign Language. Preferences also varied depending on the health topic, setting, and available technology. Importantly, participants did not expect every health resource to contain every accessibility feature. Rather, they recognized that specific accessibility features may be useful for some individuals but less useful for others, including individuals within the same sensory disability group. This suggests that accessibility does not necessarily require incorporating every possible accommodation into a single resource. Instead, participants advocated for health information that provides multiple complementary formats, allowing individuals to select the communication method that best aligns with their needs and preferences.

Our findings extend previous work by demonstrating similar variation in accessibility preferences across DHH, BLV, and DeafBlind communities, while also highlighting variation among individuals within the same sensory disability group. These findings suggest that accessibility should not be defined by a single communication modality or a checklist of individual accommodations. Rather, accessible health communication should recognize variation in communication preferences both across and within sensory disability groups and provide opportunities for individuals to choose among accessible options. This flexibility may also allow health information and recommendations to be more responsive to individual needs and preferences rather than assuming that a particular format or accommodation will work for everyone. These findings reinforce principles of universal design and person-centered communication by demonstrating that meaningful accessibility depends on providing options rather than assuming that one format will serve all users.

Participants also described trust as a prerequisite for engaging with health information. Although internet searches, artificial intelligence tools, social media, and personal testimonies were commonly used to identify information or learn from others’ experiences, healthcare providers and established medical organizations remained the most trusted sources of health information. The use of online sources in the present study aligns with prior research showing that DHH adults frequently use search engines to obtain health information (Chandanabhumma et al., 2024). Prior work has also demonstrated the value DHH adults place on experiential information, particularly information that provides insight into others’ health experiences (Chandanabhumma et al., 2024). Rather than relying exclusively on a single resource, participants frequently described verifying information across multiple trusted sources before making health decisions. This pattern indicates that people with sensory disabilities evaluate the credibility of health information and may benefit from resources that clearly identify evidence-based sources and encourage informed decision-making. As digital health information continues to expand, including the increasing availability of artificial intelligence tools, supporting individuals in identifying trustworthy information will remain an important component of accessible health communication.

Collectively, these findings highlight the importance of involving people with sensory disabilities throughout the development of accessible health information. Participants did not simply identify barriers; they offered practical recommendations grounded in lived experience that can inform future health communication efforts. Key participant-informed recommendations for improving the accessibility of health information are summarized in **Table 4**. Engaging intended users throughout the design process may help ensure that health information is not only technically accessible but also understandable, trustworthy, and responsive to the diverse communication needs of people with sensory disabilities (Oswal & Oswal, 2021). Altogether, this supports a flexible, person-centered approach to accessible health communication that combines intentional design with multiple communication options and opportunities for individual choice.

**Table 4.** Participant-informed recommendations for accessible health information.

| Recommendation | Participant-informed Strategies | Examples |
|---|---|---|
| Provide information in multiple accessible formats | Make the same health information available through complementary communication formats rather than relying on a single format. | Written text, video, audio, Braille, infographics, transcripts, captions, ASL, audio descriptions |
| Support individual choice and flexibility | Recognize that accessibility needs and communication preferences vary across and within sensory disability groups and may differ depending on the situation. Ask individuals about their preferences rather than assuming their needs. | Provide multiple format options; ask about communication preferences; allow individuals to select the option that best meets their needs |
| Use clear and concise language | Communicate health information in ways that reduce cognitive burden while maintaining the information needed to make health decisions. | Limit unnecessary medical jargon; define unfamiliar terms; use plain language and concise explanations; present key information first with additional detail available |
| Design for visual readability | Present written and visual information in ways that support readability for people with varying levels of vision. | Larger or adjustable fonts; readable font styles; adequate spacing; high color contrast; magnification options; accessible visual elements |
| Use clear organization and navigation | Structure resources so individuals can efficiently locate and navigate relevant information. | Descriptive headings; clearly separated sections; intuitive navigation; clean, simple layouts; limited advertisements and pop-ups |
| Ensure compatibility with assistive technologies | Design digital resources to function with technologies individuals use to access information independently. | Screen-reader compatibility; text-to-speech; screen magnification; alt text for images |
| Make multimedia content accessible | Provide multiple ways of accessing information communicated through video or other multimedia. | Captions/subtitles; transcripts; ASL; audio descriptions |
| Support access to trustworthy health information | Clearly communicate the source of health information and support opportunities to verify information through trusted sources. | Healthcare providers; established medical organizations; links or references to credible health information sources |
| Consider accessibility throughout resource development | Incorporate accessibility into the development of health information from the beginning rather than adding accommodations after resources have already been created. | Plan for multiple formats and accessibility features during development; consider different sensory and communication needs when designing resources; involve people with sensory disabilities in resource development |

Given the wide range of available health information resources, organizations should prioritize improving the accessibility of resources that are commonly used by the public, such as health information websites, educational videos, and patient portals. Prioritizing these resources may help organizations make the most effective use of available resources while improving access to information that individuals rely on to make informed health decisions. These efforts should be part of a broader commitment to ensuring that health information is accessible to individuals with diverse needs.

These findings also have implications for healthcare organizations’ legal obligations to provide accessible digital health information. Under Section 504 of the Rehabilitation Act, healthcare organizations receiving federal financial assistance are generally required to ensure that their web content and mobile applications meet established accessibility standards, including Web Content Accessibility Guidelines (WCAG) 2.1 Level AA, subject to specified exceptions (U.S. Department of Health and Human Services [HHS], n.d.). Similarly, updated regulations under Title II of the Americans with Disabilities Act (ADA) establish digital accessibility requirements for state and local government entities, including public healthcare organizations (U.S. Department of Justice [DOJ], 2024). Together, these requirements reinforce the importance of making digital health resources accessible to people with sensory disabilities. However, the diverse preferences identified in this study highlight the importance of complementing regulatory compliance with user-centered approaches that promote access to health information.

This study has several limitations. Participants were recruited primarily through community organizations, research networks, and online recruitment methods, which may have resulted in a sample with greater engagement in disability advocacy and health information seeking than the broader population of people with sensory disabilities. Educational level was not collected as part of the study, limiting our ability to examine how educational background may have influenced participants’ experiences accessing, understanding, and using health information. In addition, online recruitment of disability populations presents ongoing challenges related to fraudulent participation. During data collection, at least one participant was identified as providing fraudulent responses to be deemed eligible for a focus group. Following review by the research team, that participant’s responses were excluded from all analyses. While procedures were implemented to identify and remove fraudulent data, the increasing prevalence of bots and fraudulent participants highlights an important methodological challenge for disability health research conducted in online environments.

## CONCLUSION

This study highlights the importance of engaging people with sensory disabilities throughout the development of health information resources. Participants consistently emphasized that accessible health communication should be trustworthy, intentionally designed, and available in multiple complementary formats that support diverse communication preferences and promote independent access to information. These findings will inform the development of user-centered health promotion accessibility checklists to support more inclusive health communication for people with sensory disabilities.

## Author contribution statement

LSR: Conceptualization; Project administration; Formal analysis; Writing – original draft; Writing – review & editing.

CS: Investigation; Formal analysis; Writing – review & editing.

LP: Investigation; Project administration; Formal analysis; Writing – review & editing.

PFA: Conceptualization; Funding acquisition; Writing – review & editing.

DJ: Conceptualization; Funding acquisition; Writing – review & editing.

MMM: Conceptualization; Investigation; Formal analysis; Writing – review & editing; Funding acquisition; Supervision.

TGJ: Conceptualization; Investigation; Project administration; Formal analysis; Writing – review & editing; Funding acquisition; Supervision.

## Data Availability

The data reported in this manuscript are not available due to ethical and privacy concerns, including lack of informed consent by participants for sharing data.

## Acknowledgements

We would like to thank the members of our community advisory board for their dedication and work on this project. In addition, we would like to thank the participants in this research for sharing their perspectives to improve health information accessibility for people with sensory disabilities.

## Author’s note

We recognize there are a variety of terms used to describe people with sensory disabilities, including Deaf, deaf, hard-of-hearing, hearing impaired, late-deafened, blind, low vision, vision impaired, DeafBlind. In this article, we use the term “people with sensory disabilities” to be inclusive to this heterogenous population. We adopt the terms deaf/hard-of-hearing (DHH), blind/low vision (BLV), and DeafBlind throughout, and use both disability and person-first language.

